# Excellent negative predictive value (99.8%) of two rapid molecular COVID-19 tests compared to conventional RT-PCR for SARS-CoV-2 (COVID-19) in 2,011 tests performed in a single centre

**DOI:** 10.1101/2021.06.20.21258392

**Authors:** KW Leong, TL Law, AS Saiful, Kang, Woo, Chow, ZL Yong

## Abstract

Conventional Reverse Transcription Polymerase Chain Reaction (RT-PCR) remains the gold standard for testing SARS-CoV-2. Since their availability, two rapid molecular COVID-19 tests were performed in parallel with RT-PCR in all urgent and emergency admissions, as the negative predictive value was not yet ascertained. In this study, we present the data of 2,011 test results using either ID Now COVID-19 (Abbott) (Abbott ID NOW) or Xpert Xpress SARS-CoV-2 (Cepheid) (GeneXpert) tests comparing to conventional RT-PCR results. The negative predictive value is 99.8%(3 false negatives out of 1,964 tests) using a cut-off CT value of 40. Using a cut-off of RT-PCR CT value of 30 (predicting infectivity), the negative predictive value is reduced to 99.9% (1 out of 1,964 tests). With these results, we feel confident to recommend the immediate use of the rapid PCR tests alone and to use conventional RT-PCR for confirmation testing after.

## Introduction

Conventional RT-PCR for SARS-CoV-2 remains the gold standard to diagnose COVID-19 using three genes.^1^ In our centre, the test is not done in-house leading to delay of results of up to three days. For urgent and emergency admissions, this is not ideal as patients remain in isolation until a negative result is obtained, after which patients would be allowed into general wards. Previous studies of rapid COVID-19 tests reported negative predictive values using rather small sample sizes of less than 500^1,2,3,4,5^. With the relatively small sample sizes, our doctors did not feel confident to proceed with high risk procedures, such as surgeries and endoscopies, where exposure to oral and nasal secretions are high. Thus, rapid COVID-19 tests were conducted in parallel with RT-PCR.

## Methodology

Nasopharyngeal samples were taken at the same time as three swabs for tests. Oropharyngeal swabs were taken from patients with nasal bleed, very young children and patients with platelet counts below 20 (rare).

Two swabs were sent to an off-site laboratory for RT-PCR testing. TaqPath RT-PCR Assays from ThermoFisher were performed according to the manufacturers guidelines^6^. This detects three gene targets: ORF1ab, S gene and N gene. A CT cut-off value of 40 was used according to our national consensus for positivity.

The third swab was used for rapid COVID-19 testing. The samples were processed within one hour. All samples were kept fresh and not frozen. Initially, only the Abbott ID NOW kit was used. Later as GeneXpert kits were made available, both tests were used interchangeably. The Abbott ID NOW assays were performed according to the manufacturer’s guidelines. This test targets a portion of the RdRp gene within the SARS-CoV-2 genome^7^ and uses isothermal nucleic acid amplification of this target gene. The GeneXpert assays were also performed according to the manufacturer’s guidelines. This is a polymerase chain reaction test that targets the SARS-CoV-2 E and N2 genes^8,9^.

## Results

A total of 2,011 tests were performed from 7^th^ Jan - 5^th^ May 2021. A total of 1,506 tests were performed using Abbott ID now (7^th^ Jan - 5^th^ May 2021) and 505 tests using GeneXpert (from 29^th^ Mar - 5^th^ May 2021).

Using a CT value cut-off of 40 (conventional RT-PCR), positive and negative predictive values were calculated (following Malaysian consensus for reporting positive results).

There were 3 false negative results giving a negative predictive value of 99.8% (3/1,967). [Abbott ID NOW 99.8% (3/1,477); GeneXpert 100% (0/490).]

With a CT value cut-off of 30 (indicating infectivity), only one test using the Abbott ID NOW was considered a false negative (Fig. 2).

The positive predictive value is less robust at 72.7% with 12 false positives out of 44 positive tests. [Abbott ID NOW 65.5% (10/29 tests); GeneXpert 86.7% (2/15)]

The two false positives using GeneXpert were borderline results (Fig. 2) and can be considered as true positives with high CT values.

## Discussion

Our results (Fig 1a, b) show a strong and robust negative predictive value for both rapid tests at 99.8%. This will allow the rapid test to replace conventional RT-PCR in almost all situations, with conventional RT-PCR performed only as a confirmatory test. At the beginning of the pandemic, our turnaround time for conventional RT-PCR results were up to three days from collection. This improved to same-day results as more centres were able to perform RT-PCR and the pandemic temporarily slowed. However, as infection rates climbed again, and thus increasing testing demand, the turnaround time has increased again up to 36 hours.

**Figure 1a:**
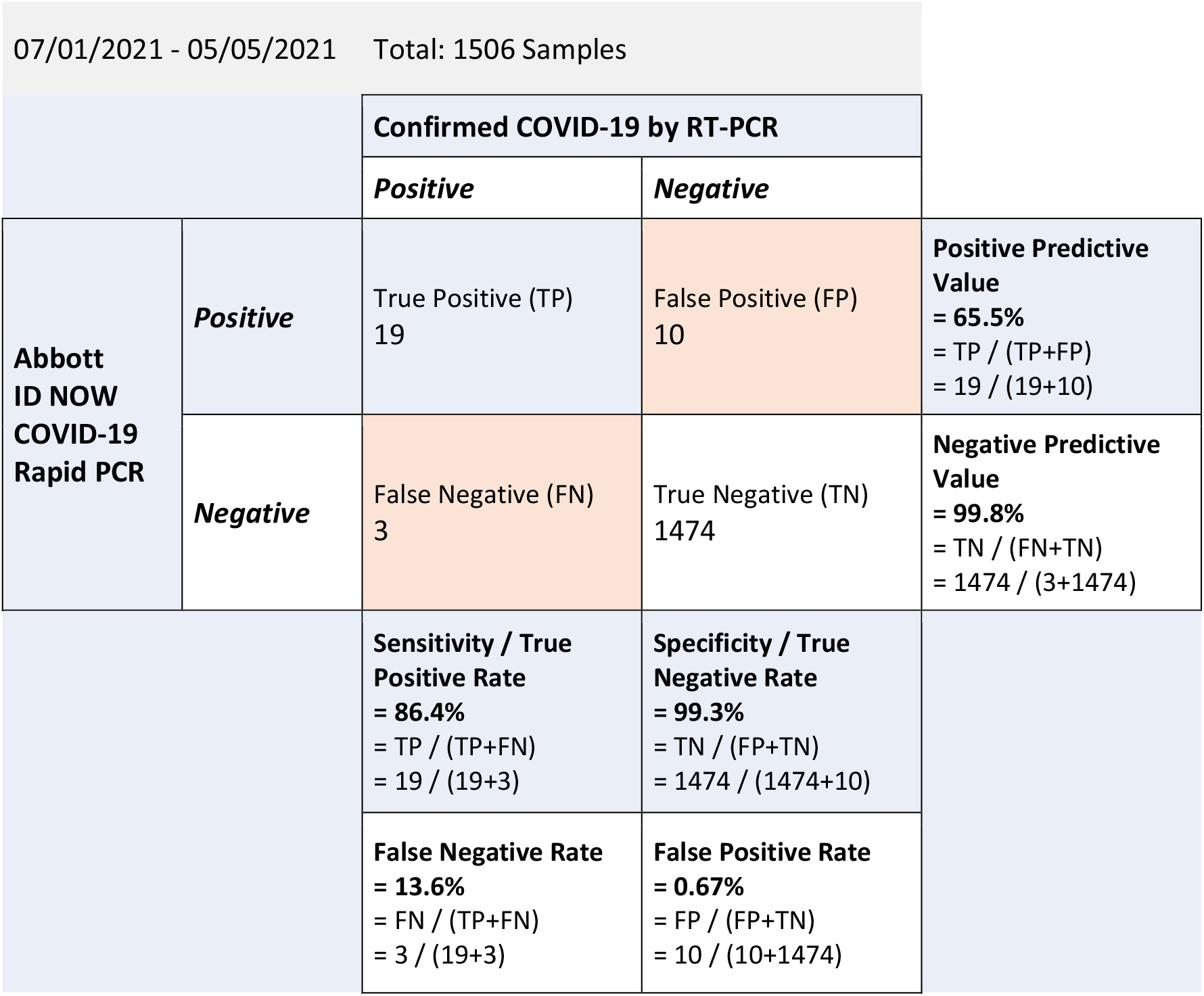
Comparison Data for Rapid Molecular Test (Abbott ID NOW) vs RT-PCR.

**Figure 1b:**
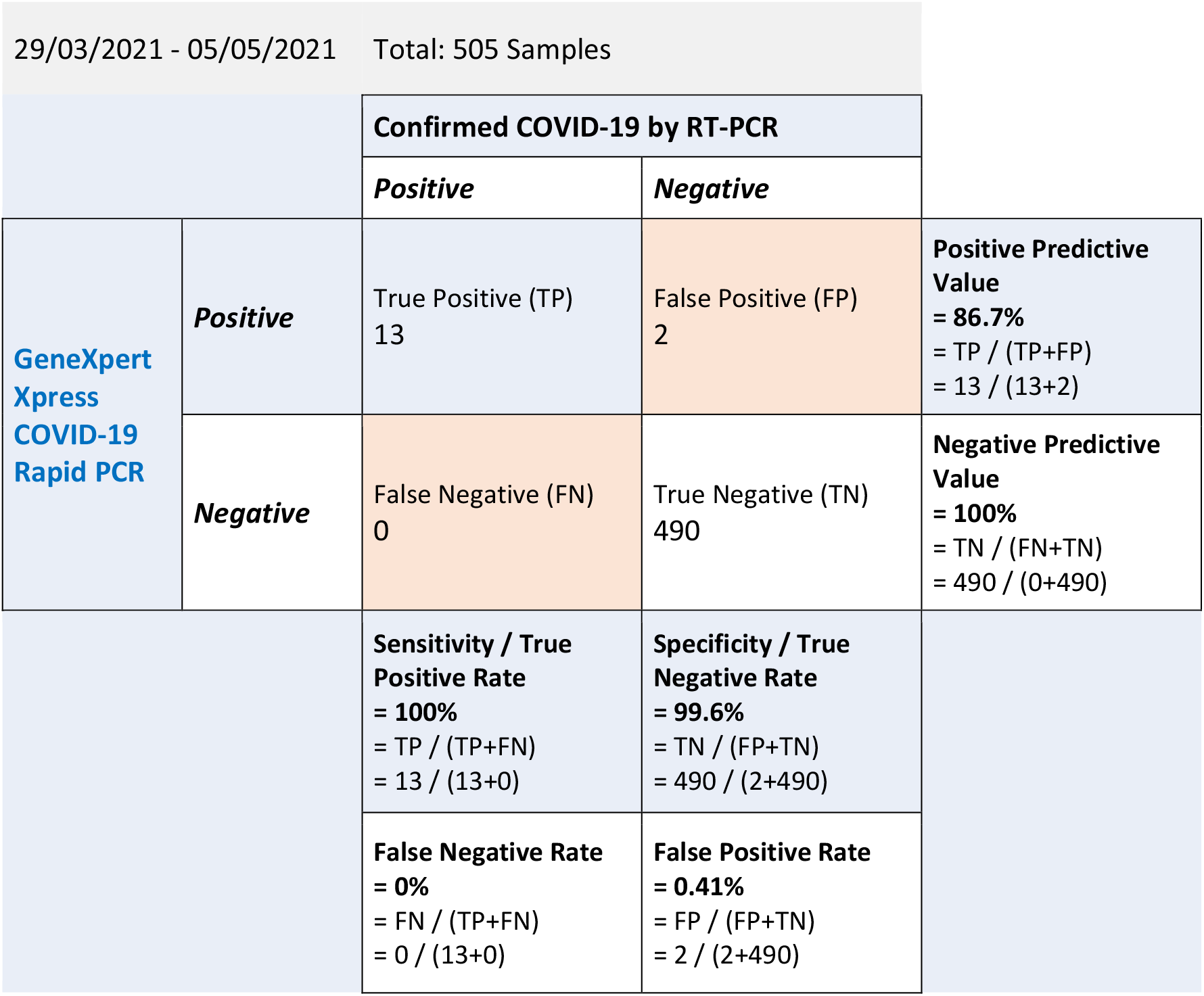
Comparison Data for Rapid Molecular Test (GeneXpert) vs RT-PCR.

Prior to the availability of these rapid COVID-19 tests, rapid antigen tests (SD Biosensor standard Q)^9^ were performed in parallel with conventional RT-PCR (3^rd^ Jun 2020 - 21^st^ Jan 2021) (Fig. 1c). The results were acceptable with negative predictive value at 99.2% (5 false negatives in 618 samples). However, there were only 6 positive cases in a total of 619 samples (0.96%) and one true positive (0.16%) corresponding to a sensitivity of 16.7% (5 false negatives of 6 positives). With these low positive results, the negative predictive value may be overestimated. This is below the sensitivity benchmark required in a hospital setting in order to continue with high risk procedures. While not ideal within the confines of a hospital, the short turnaround duration and the low expert and material requirements make the rapid antigen test a good means of community screening to reduce community transmission. Additionally, the material limiting factors (i.e., number of available tests) are not as limiting as for rapid molecular and RT-PCR tests.

**Figure 1c:**
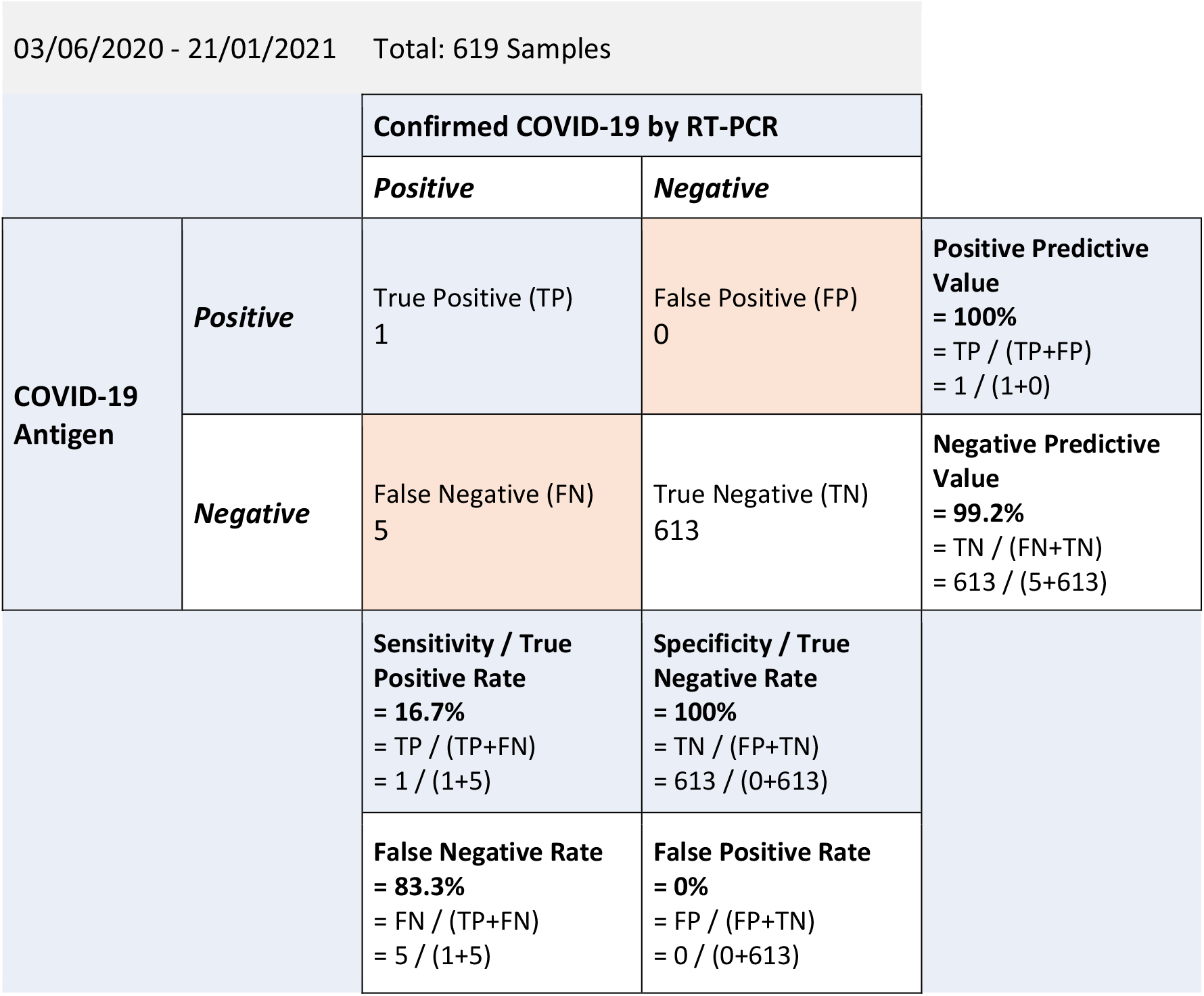
Comparison Data for COVID-19 Antigen Test vs RT-PCR.

**Figure 2a:**
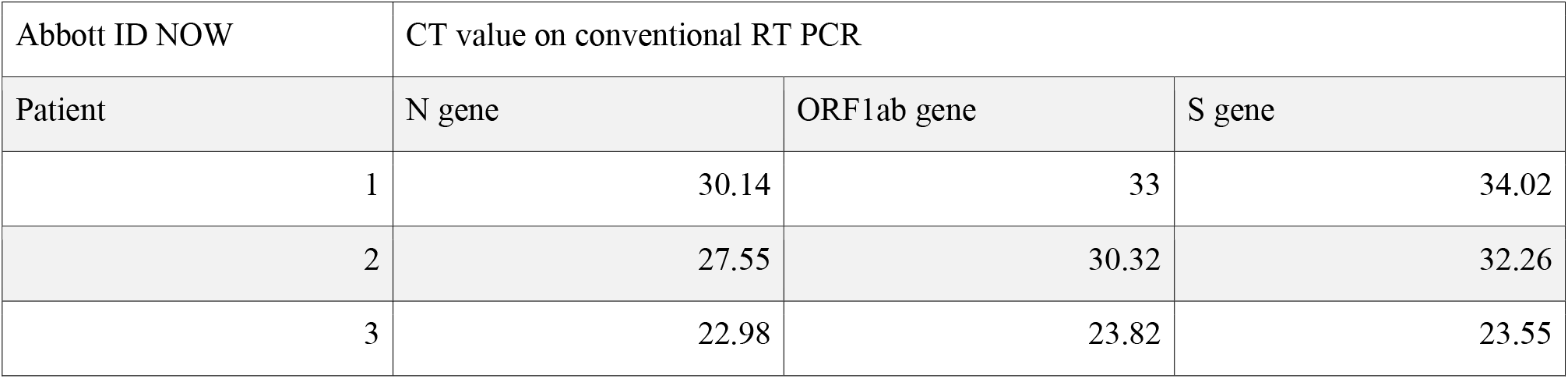
CT values for Abbott ID NOW false negatives using conventional RT-PCR.

**Figure 2b:**
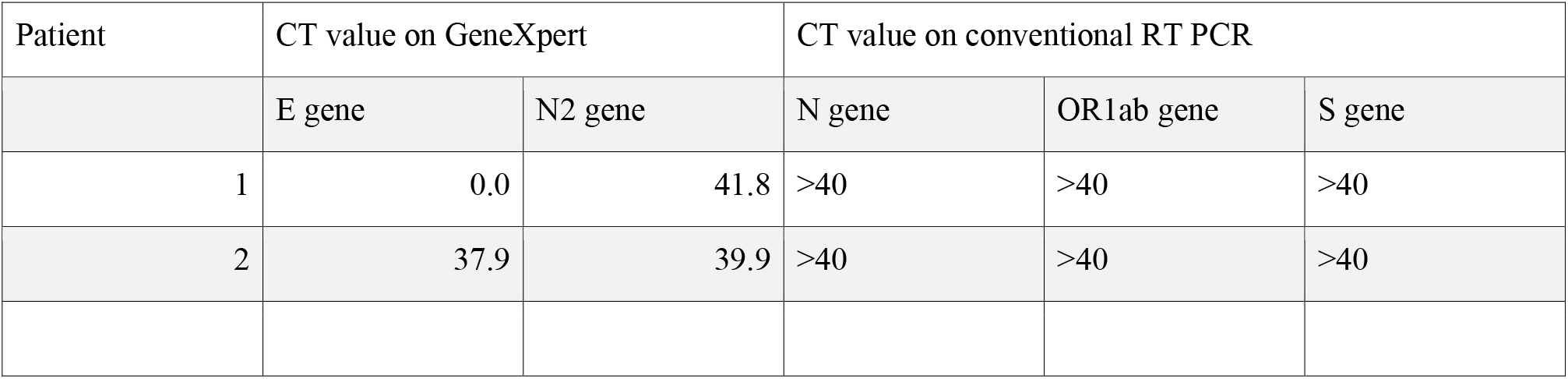
CT values for GeneXpert false positives compared to conventional RT-PCR.

Out of 2,011 tests, 32 positive tests were identified with RT-PCR (1.6% positive rate) during the period of Jan - May 2021. By contrast, there was only one RT-PCR positive test out of 619 tests during the period of Jun 2020 - Jan 2021 (0.16%). Unpublished data among the group of hospitals in May 2021 has reached 10%. In our hospital this has reached 3% in May^10^. We are currently experiencing a surge of cases since April 2021 with national rate of 5 to 6% positive tests^10^.

Both rapid molecular test kits require simple sample processing while PCR and analysis is machine automated. Turnaround time is less than one hour and the number of tests that can be performed each round is limited to the number of PCR and analysis machines. In our hospital, that number is four for each type of rapid molecular test. Consequently, as cases increase, the waiting time for a test result would increase as the number of machines becomes the limiting factor. This may be a point of concern as we face a resurgence of infections.

The positive predictive value of Abbott ID Now is 65.5% and of GeneXpert is 86.7%. Thus GeneXpert is robust and ideal point of care test. This is comparable to published results.^1,2,3,4,5^

The viral load of an infected person only increases from Day +3 of exposure and in most cases peaks on Day +5 and +8. This implies a remarkably short viral doubling time, indicating that it can take as short as two days for a person to go from non-infectious to infectious. Thus, a result may no longer be valid if the turnaround time exceeds a day.^11,12,13^ Negative conventional RT-PCR results obtained with a long turnaround time may result in false security. Thus, point-of-care testing with rapid PCR tests that take less than 1 hour is crucial when aerosolised procedures and surgeries under general anaesthesia are required. To this end, a reliable negative test for COVID-19 is essential to minimise or negate transmission of COVID-19 during procedures.

Although the positive predictive value is 65.5% for Abbott ID Now, we are reasonably satisfied with its use for primary screening. The Abbott ID Now test is likely set up to have a high negative predictive value and a corresponding higher CT value as a cutoff in order avoid missing positive cases (CT value above 35). The test utilises a single gene portion as compared to two for GeneXpert and three genes in conventional RT-PCR.

The GeneXpert returns a CT value and can measure a CT value up to 45. For comparative purposes, the CT cut off in most Malaysian laboratories is set to 40 by consensus, similar to conventional RT-PCR. In this regard, the two false positives in this report could have been true positives with low viral load as their CT value was near 40. While the RT-PCR results from these samples returned negative, the fact that the assay was carried out off site may have led to minor variations due to sample degradation over time.

Our results represent a large sample size from a single institution. Pre-analytical variables are reduced, such as sample collection and delivery. As the test swabs for both rapid tests and RT-PCR were taken at the same time, this important pre-analytical variable is further reduced. In many reported series, the rapid molecular tests were not done in parallel.

It is tempting to utilise GeneXpert without RT-PCR confirmation as it returns a CT value. However, with the rapidly changing situation and emergence of new variants, caution has to be considered in regards to how this would affect test results. We will continue monitoring the tests.

We have recommended to discontinue the use of conventional RT-PCR for routine screening of patients admitted to our hospital and reserve it as a confirmatory test. Both rapid tests are robust as screening tests. GeneXpert would be preferable with its high positive predictive value prior to high risk procedures such as intubation, upper endoscopies and aerosolised procedures. Being point-of-care tests, the tests can easily be repeated should procedures be delayed. However, the number of tests performed per time would be a limiting factor. The Abbott ID NOW is a robust test to complement GeneXpert.

## Data Availability

All data are kept in the laboratory computer system.

